# Basrah experience among 6404 patients with COVID-19

**DOI:** 10.1101/2020.10.19.20215384

**Authors:** Saad S. Hamadi Al-Taher, Abbas K AlKanan, Mohammad N. Fares, Nihad Q. Mohammed, Ali Raheem Al-Jabery, Awatif A. Habeeb, Abbas Ali Mansour

## Abstract

**Background:** The first case of COVID-19 report in Basrah was in early March 2020. This study aimed to assess some of the characteristics of patients with COVID-19 in Basrah for the period from March, 4^th^ to September, 8^th^ 2020.

**Methods:** Retrospective database analysis of the University of Basrah database. All RT-PCR positive patients during the study period were enrolled.

**Results:** Of 6404 patients included, male constituted 54.8%. Healthcare workers constituted 11.4% of the infected people. Of health care workers 16.1% were physicians. The mean age for the whole cohort was 39±16.7 years; adolescents and children younger than 20 years constituted 12.4%. The peak age was 31-40 years, those aged 61 years or more constituted 9.8% only. The case fatality rate was 3% (males 55.2% and females 44.8%). No death was reported in adolescents or children. The highest death rate was among those age 61 years or more.

**Conclusion:** The situation of COVID-19 infection in Basrah, Iraq is evolving like other countries. Furthers studies are needed to assess associated comorbidities, treatment lines, outcomes and variables associated with mortality.

---

March 11, 2020 the World Health Organization established COVID-19 as pandemic after its first appearance in Wuhan, China in December 2019. This pandemic uncovered major threats to the health system of all countries. Clearly, it shows how fragile the health structure worldwide. The high number of healthcare workers infected till now despite all the recommendation of personal protections uncovers disastrous gaps in the knowledge on the disease. The care for patients with noncommunicable diseases faced tragic situation because of shift of almost all care toward patients with COVID-19.(1)

The social distancing causes an unpreceded direct and indirect psychological trauma including the direct effect on economy.(2)

Telemedicine and teaching realignments in all school are the main global changes seen.

Paucity of symptoms, wide range of incubation period (2-14 days) and false negative reverse transcriptase PCR (RT-PCR) test were major obstacles to contain the outbreak by imposing quarantine.(3)

The first case report of COVID-19 in Iraq was in the Najaf city of an Iranian student on February 24. The city of Basrah, in the south of Iraq, reported the first case on March 9 and the first death in March 10.(4)

By the end of February 2020, the health authority of Basrah (Basrah Health Directorate) in collaboration with University of Basrah started to modify the local health system in the city to accommodate for this new disease including preparing one major hospital (Basrah Teaching Hospital) to receive COVID-19 cases, thereafter more than 5 hospital become equipped to deal with patients infected with COVID-19.

By the mid-June, it was decided to resume the care for non-COVID-19 patients because the number of cases from February to mid -June was not very high (around 10-20 cases daily). Unfortunately, by mid-June there was sudden increase in the number of newly discovered cases up to 250 -350 daily. During that period there were periods of alternating lockdown and relaxation until 24th of September, when the lockdown was totally lifted.

In the period from February to mid-June 2020, almost all COVID-19 cases were obliged by health authority to be hospitalized for at least 10 days and they need to be twice RT-PCR negative before discharge. Thereafter home isolation became the standard because of the dominance of mild cases.

Noteworthy, diabetes mellitus, specially that requiring insulin use is considered a strong risk factor for progression of COVID-19. Iraq is one of highest prevalence countries for diabetes (reaching up to 20% in Basrah). This could explain the increased number of severe cases in Basrah.(5)

This study aimed to assess some of the characteristics of patients with COVID-19 in Basrah for the period from March, 4th to September, 8th 2020.

## Methods

### Designs

Retrospective database analysis of the University of Basrah database was done for the period March, 4^th^ to September, 8^th^ 2020. The study was approved by the ethical committee of Basrah University.

### Precipitants

All RT-PCR positive patients during this period were enrolled. Data collected by the University of Basrah.

The following patients were excluded: Negative chest computerized tomography (CT) signs and negative RT-PCR, those who died at home with no known RT-PCR status or those treated in the private sector based on inflammatory markers and or CT chest with no know RT-PCR status.

Variables assessed -are: gender, job, residency, age distribution according to gender and age stratum. Case fatality rate and the mean age group death.

### Laboratory analysis and imaging

In the period from March to end of April the samples were send to the ministry of health in Baghdad to perform the RT-PCR. At the end of April 2020 many local laboratories were established in in Basrah providing RT-PCR results within 48 hours using the WHO standard.

### Data analyses

Data expressed as number and percentage of mean ±SD accordingly.

## Results

Of 6404 study cohort involved, male (3489/6404) constituted 54.8%; 33.6% (2154/6404) were employed and (2194/2915) 75.3% of women were housewives. Healthcare workers involved in 11.4% (733/6404) of whole study cohort. Of 733 health care workers 118(16.1%) were doctors and 615(83.9%) were other health care workers. About 63% (4035/6404) from peripheral city districts (Table-1).

**Table-1.**
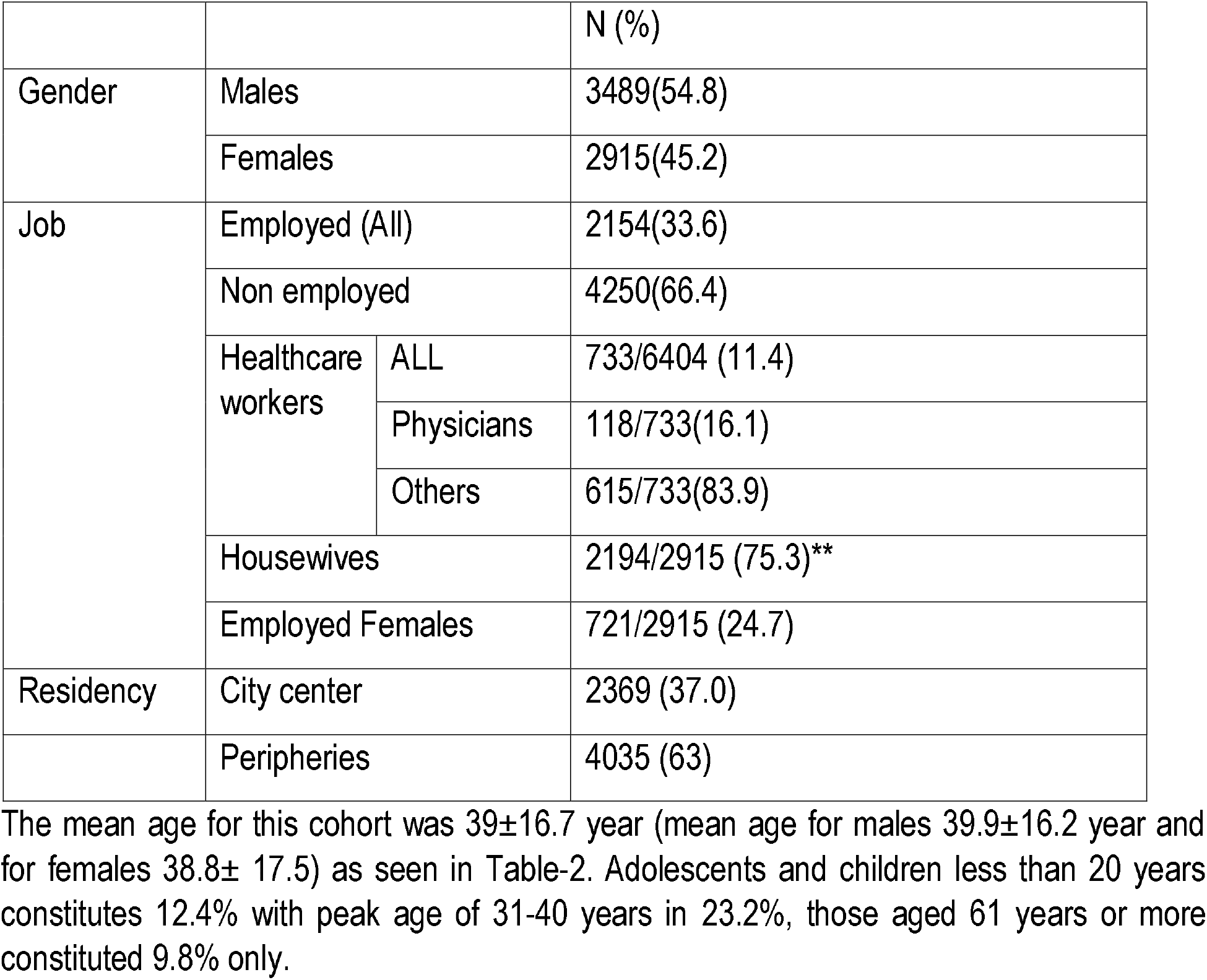
Basic demographic data of 6404 patients with COVID-19 in Basrah.

**Table-2.** Distributions of 6404 patients with COVID-19 in Basrah according to age group.

|  |  |  |
| --- | --- | --- |
| | | N (%) or mean ±SD |
| Mean age years | Mean ±SD | $39\pm 16.7$ |
| Mean age males | Mean ±SD | $39.9\pm 16.2$ |
| Mean age females | Mean ±SD | $38.8\pm 17.5$ |
| Age distribution | Less than 20 years | 792 (12.4) |
|  | 21-30 | 1336 (20.8) |
|  | 31-40 | 1336 (23.2) |
|  | 41-50 | 1314 (20.5) |
|  | 51-60 | 894 (13.96) |
|  | 61 or more | 631 (9.8) |

**Table-3.**
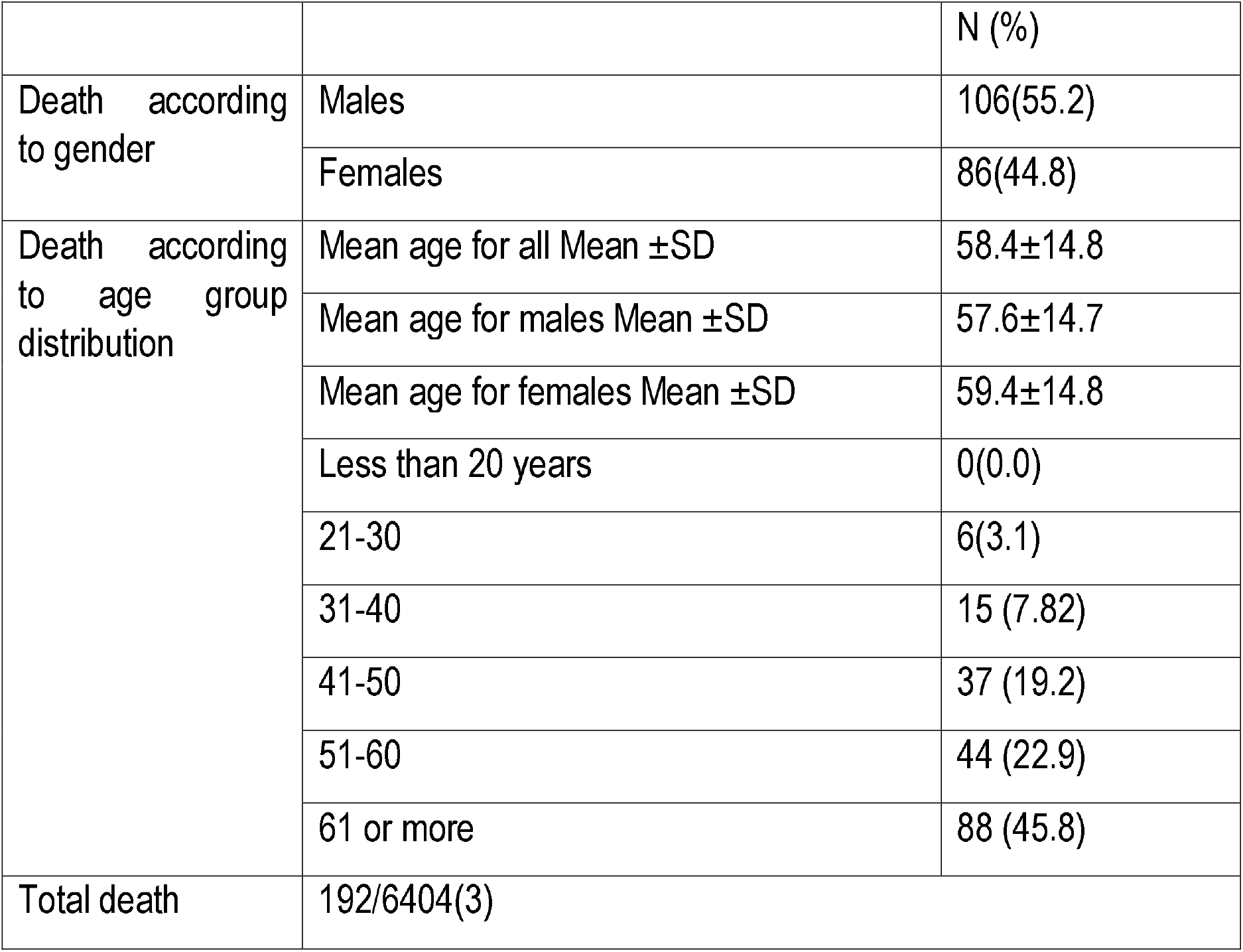
Distributions of death among 6404 patients with COVID-19 in Basrah.

## Discussion

Underdoing RT-PCR is problem all over the worlds and only one-tenth of symptomatic patients with serology positive reported previous nasopharyngeal swabs done.(3, 6)

Furthermore, 40% of COVID-19 may be asymptomatic making the true incidence and prevalence almost impossible to be predicted by any study as is the mortality rate.(7)

Published data on COVID-19 in Iraq are among the lowest all over the worlds. (8-12)

Males slightly affected more than females in more than halve in this study. The same gender prevalence was reported, unlike that of Saudi Arabia were males contributed to 80% of cases.(13, 14)

Healthcare affected is around one-tenth of the total infected patients. Worldwide, among 2,035,395 community individuals there were 99,795 healthcare workers infected (adjusted HR 11·61, 95% CI 10·93–12·33).(15).

One-tenth of cases were adolescent and children and the peak age of affected patients was 31-40 years in this study. In Saudi Arabia 90% of cases were adults and 10% only children and elderly, while in the western countries, the peak age is 20-29 years.(14, 16)

There were more deaths in males compared to women. Furthermore, around halve of deaths were are in those aged 61 years or older with case fatality rate of 3% in our cohort. The case fatality was very low in Saudi Arabia of around 0.2% and in outher countries ranging 2.7-7%% according to the disease waves.(17, 18)

### Study limitation

We think that the prevalence of COVID-19 infection is underestimated due to multiple factors, such as: Majority of people with symptoms compatible with COVID-19, do not undergo RT-PCR testing, majority of mild to moderate cases were treated in the private sector without RT-PCR testing. Beside that a lot of death at home were not labeled as COVID-19 because no tests have been done. Comorbidities and smoking status were not studied in this report and it will the project for future work.

## Data Availability

All data available

## Conclusion

Compared with COVID-19 infection in Basrah, Iraq to other countries, Basrah experienced similar burden of disease with comparable outcome. Further studies are needed to assess the comorbidities associations, treatment lines and outcome.

## Competition of interest

None

## Authors contribution

All authors contributed equally to this work.

